# Angiotensin II receptor I auto-antibodies following SARS-CoV-2 infection

**DOI:** 10.1101/2021.06.30.21259796

**Authors:** Whitney E. Harrington, Yonghou Jiang, Fergal Duffy, Jennifer Hadlock, Andrew Raappana, Sheila Styrchak, Ingrid Beck, William Chour, John Houck, Venkata Duvvuri, Winnie Yeung, Micaela Haglund, Jackson Wallner, Julie A. Wallick, Samantha Hardy, Alyssa Oldroyd, Daisy Ko, Ana Gervassi, Kim M. Murray, Henry Kaplan, John D. Aitchison, James R. Heath, D. Noah Sather, Jason D. Goldman, Lisa Frenkel

## Abstract

**BACKGROUND:** Coronavirus disease 2019 (**COVID-19**) is associated with endothelial activation and coagulopathy, which may be related to pre-existing or infection-induced pro-thrombotic autoantibodies such as those targeting angiotensin II type I receptor (**AT1R-Ab**).

**METHODS:** We compared prevalence and levels of AT1R-Ab in COVID-19 cases with mild or severe disease to age and sex matched negative controls.

**RESULTS:** There were no significant differences between cases and controls. However, there were trends toward a higher proportion with AT1R-Ab positivity among severe cases versus controls (32% vs. 11%, p=0.1) and higher levels in those with mild COVID-19 compared to controls (median 9.5U/mL vs. 5.9U/mL, p=0.06).

**CONCLUSIONS:** These findings suggest that AT1R-Ab are not consistently associated with COVID-19 but do not exclude a contribution to endothelial pathology in a subset of people.

## Introduction

Coagulopathy is emblematic of severe disease from severe acute respiratory syndrome coronavirus 2 (**SARS-CoV-2**) infection (1). Series from patients admitted to the intensive care unit and autopsy series describe high rates of both pulmonary embolism and small vessel inflammation and thrombosis in the lungs and other organs (2, 3). A number of mechanisms that may mediate coronavirus disease 2019 (**COVID-19**)-associated coagulopathy have been proposed including direct endovascular damage, altered platelet function, and pre-existing or infection-induced pro-thrombotic auto-antibodies (1). Pro-thrombotic anti-phospholipid antibodies have been described in some but not all patients with COVID-19 associated coagulopathy (4), and recent research suggests the potential for cross-reactive auto-antibodies following infection with as yet unidentified targets (5).

Amongst previously described anti-endothelial antibodies, those directed against the angiotensin II type 1 receptor (**AT1R-Ab**) are associated with essential hypertension (6), pre-eclampsia (7), and vascular rejection following renal transplant (8). AT1R is part of an angiotensin II-driven signaling cascade that leads to increased blood pressure and inflammatory cytokine production; the pathway may be inhibited by the activity of ACE2, which interestingly also functions as the primary receptor for SARS-CoV-2 cell entry (9). In particular, AT1R-activating auto-antibodies (**AT1R-AA**) directed against the second extracellular loop of AT1R are associated with pathology (7, 8), including elevated pro-inflammatory TNF-a and IL-6 cytokine levels and increased disease severity in pre-eclampsia models (7). AT1R-AA are reported to induce expression of tissue factor by vascular smooth muscle which may trigger aberrant coagulation and clot formation (10). In renal transplant recipients, AT1R-AA are associated with refractory rejection and malignant hypertension, as well as vasculopathy including arterial inflammation, endothelial activation, tissue factor expression, and thrombosis (8).

Multi-organ endothelial inflammation is strongly associated with disease severity and poor outcome from COVID-19 (2, 3, 11). Such endothelial damage may allow the binding of or trigger the development of anti-endothelial antibodies such as AT1R-Ab. Alternatively, structural homology between epitopes of the SARS-CoV-2 Spike and the second extracellular loop of AT1R might lead to the development of cross-reactive antibodies. These possibilities led us to consider AT1R-Ab as a potential mediator of COVID-19-associated coagulopathy and disease. If directed against the second extracellular domain of AT1R, such antibodies might trigger hypertension, inflammation including cytokine storm, and pulmonary edema as seen in severe COVID-19. In addition, these antibodies might contribute to further endothelial inflammation, tissue factor expression, and hypercoagulability. We therefore tested the hypothesis that COVID-19 infection is associated with a higher prevalence and levels of AT1R-Ab and assessed whether SARS-CoV-2 negative individuals with AT1R-Ab demonstrated cross-reactivity against SARS-CoV-2 Spike trimer.

## Materials & Methods

### Cohorts and samples

Mild COVID-19 cases were from the Seattle Children’s SARS-CoV-2 Recovered Cohort and the Seattle Children’s SARS-CoV-2 Prospective Cohort both approved by Seattle Children’s Hospital IRB, with study specimens from ≥14 days from COVID-19 symptom onset (**Table 1**). Participants recovered at home, except for one participant who required hospitalization for oxygen therapy but no additional support. Severe COVID-19 cases were participants in the Swedish-Institute for Systems Biology Novel Coronavirus (**INCOV**) Biobank, approved by Providence St. Joseph Health IRB, with samples from ≥7 days from symptom onset. The severe COVID-19 cases had maximum WHO COVID-19 scores between 4 and 8 (median 5). Six of 19 participants with severe disease died from complications of COVID-19. Human leukocyte antigen (**HLA**) typing by direct sequencing had previously been conducted for the INCOV cohort. Cases were matched 1:1 to controls by age and sex. Healthy controls were derived from the Children’s SARS-CoV-2 Prospective Cohort. At the time of blood collection (March to July, 2020), controls reported no prior history of SARS-CoV-2-like illness, were nasal specimen PCR negative for SARS-CoV-2, and were SARS-CoV-2 antibody negative on the SCoV-2 Detect™ IgG and IgM ELISAs (InBios, Seattle, WA). All participants provided written informed consent.

**Table 1.** Cohort demographics by COVID-19 status.

|  | All COVID-19 Controls & Cases |  |  | Mild COVID-19 Controls & Cases |  |  | Severe COVID-19 Controls & Cases |  |  |
| --- | --- | --- | --- | --- | --- | --- | --- | --- | --- |
| Demographics | Controls | Cases | p-value | Controls | Cases | p-value | Controls | Cases | p-value |
| Number | 53 | 53 |  | 34 | 34 |  | 19 | 19 |  |
| Age, years, median [range] | 46 [24-73] | 45 [24-88] | 0.8 | 41 [24-71] | 40 [24-74] | 1 | 62 [38-73] | 64 [38-88] | 0.6 |
| Female, n (%) | 34 (64%) | 34 (64%) | 1 | 27 (79%) | 27 (79%) | 1 | 7 (37%) | 7 (37%) | 1 |
| Days from 1st symptom, median [range] |  | 24 [7-82] |  |  | 34 [14-82] |  |  | 13 (7-37) |  |

### Structural Modeling

To assess the biologic plausibility of cross-reactive antibody binding sites, searches to find potential regions of structural homology between AT1R and SARS-CoV-2 Spike protein were performed. The UCSF Chimera MatchMaker tool was used, which aligns structures based on pairwise sequence alignment followed by 3D structural alignment using residue Cα positions. Spike protein monomer [PDB: 6VXX] was used as the SARS-CoV2 reference structure compared to human AT1R protein [PDB: 4YAY].

### Anti-AT1R Antibody Screening

The concentration of AT1R-Ab in plasma was measured with a quantitative ELISA (Celltrend, Luckenwalde, Germany) using the entire AT1R protein, with assays performed according to the manufacturer’s instructions. In brief, standards and diluted samples (1:100) were added to AT1R-precoated microtiter plates and incubated for 2 h at 4°C. After washing, AT1R-Ab was detected with HRP-labelled anti-human IgG antibody followed by enzymatic substrate reaction. Samples were tested in duplicate. Optical densities (**OD**) were converted into concentrations by comparison to a standard curve and mean concentration was used to define a sample as positive when >=17 U/mL, indeterminate when 10–16.9 U/mL, and negative when <10 U/mL according to the manufacturer’s recommendation.

### Anti-SARS-CoV-2 Spike, Receptor Binding Domain, and Nucleocapsid Protein ELISAs

In order to investigate potential cross-reactivity between AT1R-Ab and SARS-CoV-2 Spike, we tested plasma from AT1R-Ab positive control subjects and randomly selected AT1R-Ab negative controls for reactivity against SARS-CoV-2 Spike trimer (including both S1 and S2 domains), as previously published (12). Because we hypothesized that potential cross-reactive antibodies might bind SARS-CoV-2 Spike with lower avidity, we started with an initial plasma dilution of 1:10 followed by serial dilutions by a factor of two. To assess for potential cross-reactivity induced by prior non-SARS-CoV-2 coronavirus infection, we additionally tested these control subjects against the receptor binding domain (**RBD**) in S1 and the nucleocapsid protein (**NP**) using a similar ELISA format. Endpoint titers were defined as the reciprocal of plasma dilution at OD 0.1 at a wavelength of 450nm after the subtraction of plate background. Positive antibody responses were defined at a titer greater than or equal to 1:50 for each antigen.

### Statistical Analysis

AT1R-Ab prevalence and levels in all COVID-19 cases were first compared to those in the entire set of controls. In secondary analysis, the subsets with mild and severe COVID-19 were separately compared to their age and sex matched controls. The distribution of AT1R-Ab status (positive versus intermediate/negative) was compared between groups using the Chi-square test, and AT1R-Ab level was compared between groups using quantile regression on the median. Distribution of HLA and SARS-CoV-2 antibody responses were compared with the Chi-square test. Significance was defined as p≤0.05, whereas trend was defined as p≤0.1.

## Results

### Association between AT1R-Ab and SARS-CoV-2 infection

Participants with mild versus severe COVID-19 were younger and more likely to be female (**Table 1**), consistent with prior reports of older age and male sex as risk factors for severe COVID-19.

There was no significant difference in the proportion of AT1R-Ab positive participants between cases and controls (26% vs. 17%, p=0.3) or in the subset of mild cases versus controls (21% vs. 21%, p=1) (**Figure 1A, 1B**). However, there was a trend toward higher proportion with AT1R-Ab positivity among severe cases versus controls (32% vs. 11%, p=0.1) (**Figure 1C**). Similarly, there was not a statistically significant difference in the median AT1R-Ab level between any group, but a trend toward higher AT1R-Ab levels was observed in all cases versus controls (median 9.0 vs. 6.0 U/mL, p=0.1), which was also found in the subset of mild COVID-19 cases and controls (median 9.5 vs. 5.9 U/mL, p=0.06), but not severe COVID-19 cases and controls (both medians 6.7 U/mL, p=1) (**Figure 1**).

**Figure 1.**
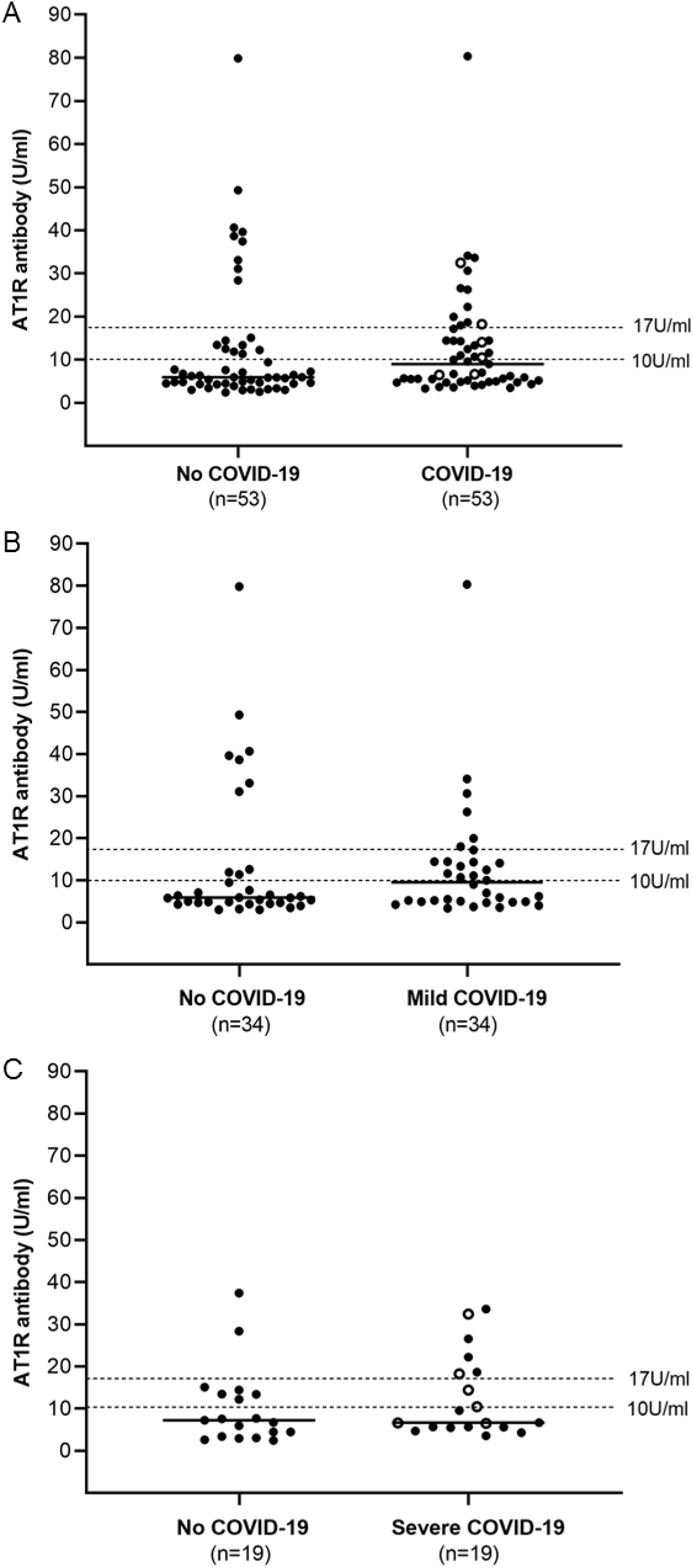
Association between AT1R-Ab and COVID-19. AT1R-Ab levels < 10U/mL were defined as negative, 10-16.9 indeterminate, >= 17 positive (dotted lines). Solid lines indicate medians. Open circles are cases with known thrombosis. (**A**) There was a trend toward higher level of AT1R-Ab in all cases versus controls (median 9.0 vs 6.0, p=0.1) and (**B**) amongst the subset of mild COVID-19 versus controls (median 9.5 vs 5.9, p=0.06). (**C**) There was a trend toward higher proportion positive for AT1R-Ab amongst severe cases versus controls (32% versus 11%, p=0.1).

### Association between AT1R-Ab, thrombotic events, and HLA-type amongst severe COVID

Within the severe COVID-19 group, we tallied six participants with thrombotic events. These included one participant with a myocardial infarction and one with a pulmonary embolism, both with high levels of AT1R-Ab (33 and 18 U/mL respectively), and four participants with deep venous thrombosis, with two having indeterminant levels of AT1R-Ab (11 and 14 U/mL). The proportions of AT1R-Ab positive participants were not different in those with and without thrombosis (33% vs 31%, p=0.9). While the median level of AT1R-Ab was higher in those with thrombosis, this was not statistically significant (12.5 vs. 5.7 U/mL, p=0.3).

AT1R-Ab have previously been associated with the HLA DR1*04 group (13). A higher proportion of AT1R-Ab positive versus negative participants carried a DRB1*04 allele; however, this was not statistically significant (50% vs. 18%, p=0.2) (**Supplementary Table 1**).

### Structural homology between AT1R and SARS-CoV-2 Spike and assessment of antibody cross-reactivity

Potential cross-reactivity of antibodies between SARS-CoV-2 Spike and AT1R was predicted based on our finding of structural homology between the aligned proteins in the S2 domain of the SARS-CoV-2 Spike and the six-membered alpha-helical bundle of AT1R, including the second extracellular domain targeted by AT1R-AA (**Figure 2**). To test the hypothesis that AT1R-Ab cross-reacts against SARS-CoV-2 Spike, in particular the S2 domain, the nine AT1R-Ab positive SARS-CoV-2 uninfected control participants and six randomly selected AT1R-Ab negative SARS-CoV-2 uninfected control participants were tested for reactivity against SARS-CoV-2 Spike trimer by ELISA. AT1R-Ab status was not associated with reactivity to SARS-CoV-2 Spike trimer (17% AT1R-Ab negative vs. 22% positive control participants, p=0.8). In addition, some AT1R-Ab positive control participants demonstrated low level reactivity against RBD (0% AT1R-Ab negative vs. 22% positive participants, p=0.2) and NP (17% AT1R-Ab negative vs. 44% positive participants, p=0.3) (**Figure 3**).

**Figure 2.**
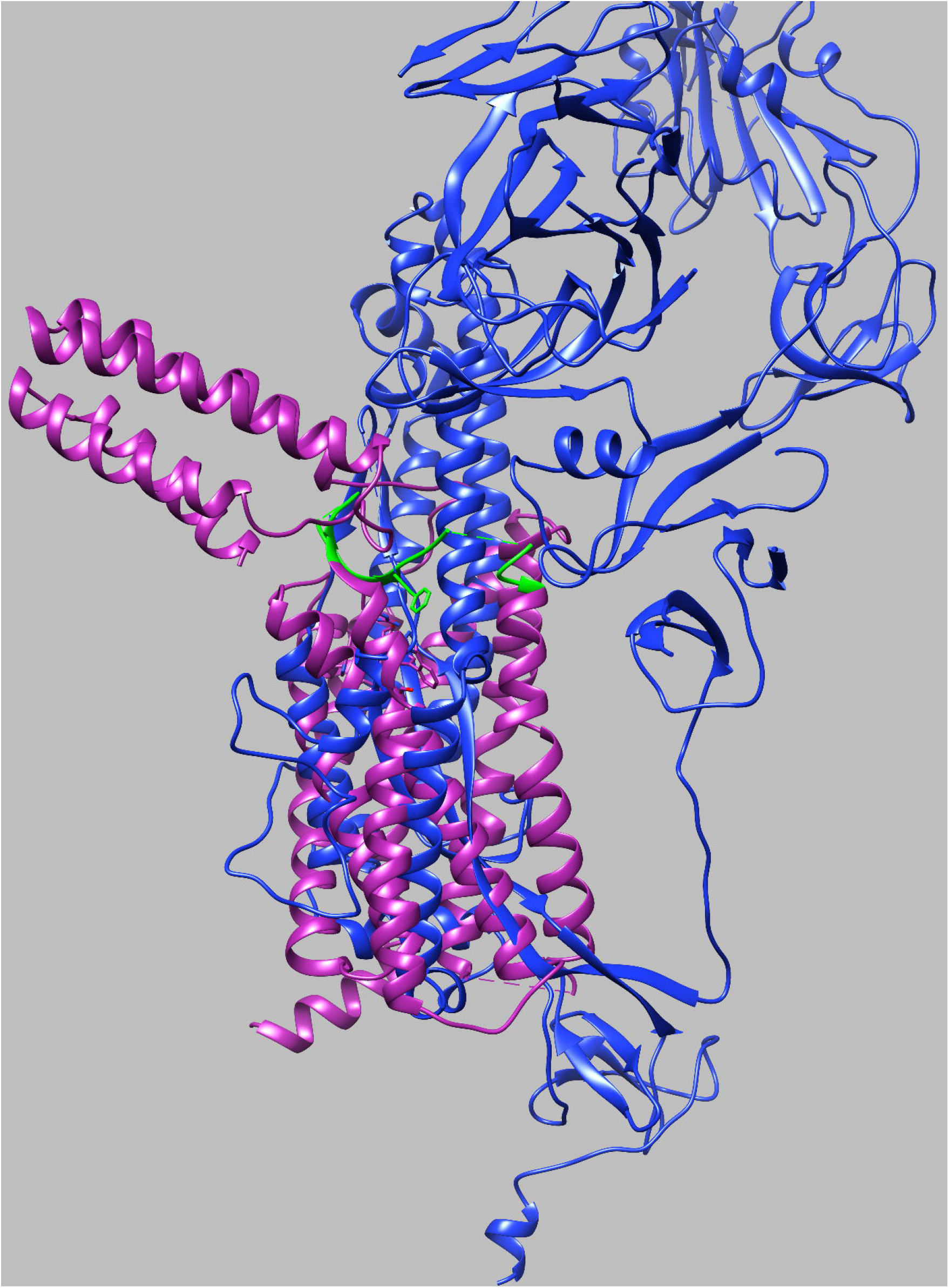
Structural overlap between SARS-CoV-2 Spike and AT1R. The structures of SARS-CoV-2 Spike (S1 and S2) and AT1R were aligned to assess potential homology. Overlap was identified between SARS-CoV-2 Spike (blue) in the S2 domain with the six-membered alpha-helical bundle of AT1R (purple), including the second extracellular domain (green). The root-mean square deviation over all 237 aligned residue pairs was 35.6 Angstroms.

**Figure 3.**
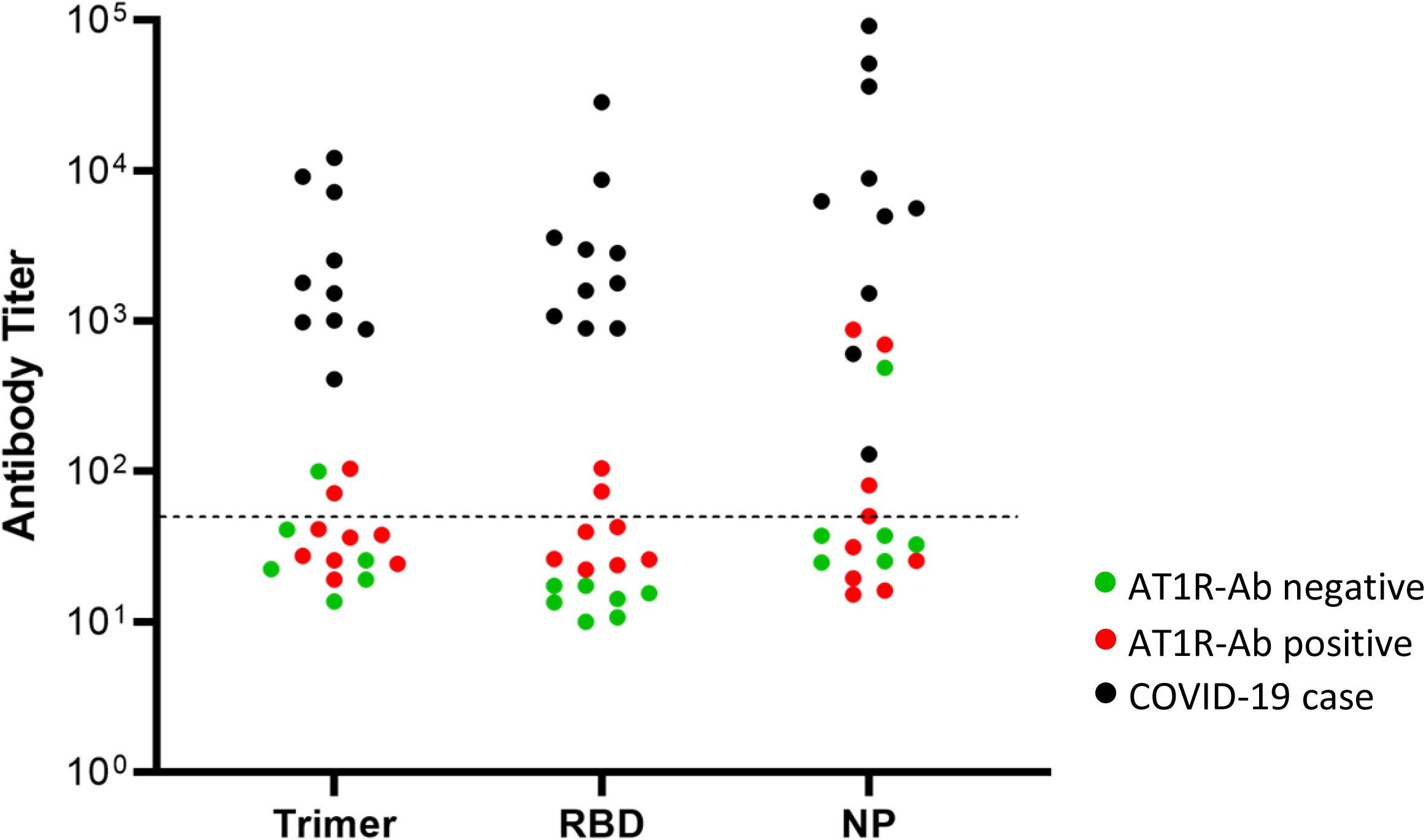
Assessment of SARS-CoV-2 protein immune reactivity by AT1R antibody status. ELISA for SARS-CoV-2 Spike trimer, RBD, and NP. Green: AT1R Ab negative, SARS-CoV-2 negative controls. Red: AT1R Ab positive, SARS-CoV-2 negative controls. Black: SARS-CoV-2 positive cases. Endpoint titers were defined as the reciprocal of plasma dilution at OD 0.1 read at 450nm after the subtraction of plate background. Positive threshold was defined as 1:50 and is indicated with dashed line. There was no clear relationship between AT1R antibody status amongst SARS-CoV-2 negative subjects and immunoreactive pattern against SARS-CoV-2 proteins.

## Discussion

Overall, AT1R-Ab levels were not significantly different between controls and cases with either mild or severe COVID-19. However multiple trends were observed. Specifically, amongst severe COVID-19 cases there was a trend towards an increase in the proportion of cases versus controls who were AT1R-Ab positive and a trend towards higher levels of AT1R-Ab amongst mild COVID-19 cases versus controls. Among severe cases, there was no significant association with level or prevalence of AT1R-Ab and a thrombotic event. Although we did not identify any significant associations, the trends we observed are consistent with a recent report of an association between AT1R-Ab and worse outcomes in patients hospitalized for COVID-19, suggesting they may play a role in endothelial activation during COVID-19 (14).

Amongst severe COVID-19 cases, where HLA typing was available, half of AT1R-Ab positive or indeterminate participants carried a DRB1*04 allele (13). Of note, DRB1*04:01 and DRB1*04:05 are both associated with increased risk of autoimmune disease (15). These participants may have had pre-existing AT1R-Ab not associated with their SARS-CoV-2 infection. Alternatively, when infected with SARS-CoV-2, individuals with this allele may have had a higher risk of developing AT1R-Ab in the setting of endothelial damage. We are not able to dissociate these possibilities given the cross-sectional nature of our sampling at the time of COVID-19.

We identified a region of potential structural homology between AT1R and the S2 domain of SARS-CoV-2 Spike; however, among controls without SARS-CoV-2 there was no clear association between AT1R-Ab reactivity and low-level reactivity against SARS-CoV-2 Spike trimer arguing against cross-reactivity as a driver of AT1R-Ab development. In addition, some controls reacted against RBD and NP, suggesting that the reactivity we detected may represent cross-reactive antibodies from prior endemic coronavirus infections (16). It is possible but less likely that these participants had experienced an unrecognized SARS-CoV-2 infection given their commercial antibody negative status and the time of enrollment when the prevalence of infection remained low in the greater Seattle area.

Our study was limited by small sample size, particularly amongst our severe COVID-19 cases. In addition, as we did not have samples from cases that pre-dated their SARS-CoV-2 infections we could not determine whether AT1R-Ab were pre-existing or developed in the setting of SARS-CoV-2 infection. Further, we were not able to determine whether AT1R-Ab positive controls are likely to develop more severe COVID-19 if SARS-CoV-2 infected.

An improved understanding of the mechanisms driving COVID-19-associated coagulopathy could point to therapies to lessen COVID-19 morbidity and mortality. We did not identify a significant association between AT1R-Ab and COVID-19 severity in this small case-control study, but the trends we observed support a possible association between AT1R-Ab and COVID-19. Further research should explore whether other endothelial autoantibodies are associated with hypercoagulability in COVID-19.

## Data Availability

Data is available upon request from Dr. Harrington

