## Supplementary Table 1 for "Angiotensin II receptor I auto-antibodies following SARS-CoV-2 infection"

**Supplementary Table 1. Human leukocyte antigen DRB1 allele by AT1R-Ab status amongst those with severe COVID-19**

| **AT1R-Ab** | **Level (U/mL)** | **Loci** | **Allele 1** | **Allele 2** |
| --- | --- | --- | --- | --- |
| Positive | 33.6 | DRB1 | DRB1*15:02:01 | DRB1*15:02:01 |
| Positive | 32.5 | DRB1 | DRB1*04:05:01 | DRB1*15:02:01 |
| Positive | 26.6 | DRB1 | DRB1*14:04:01 | DRB1*15:02:01 |
| Positive | 22.2 | DRB1 | DRB1*04:05:01 | DRB1*15:02:01 |
| Positive | 18.7 | DRB1 | DRB1*09:01:02 | DRB1*11:01:01 |
| Positive | 18.2 | DRB1 | DRB1*04:01:01 | DRB1*13:02:01 |
| Indeterminate | 14.5 | DRB1 | DRB1*03:01:01 | DRB1*12:01:01 |
| Indeterminate | 10.5 | DRB1 | DRB1*04:04:01 | DRB1*04:05:01 |
| Negative | 9.6 | DRB1 | DRB1*04:01:01 | DRB1*14:216 |
| Negative | 6.7 | DRB1 | DRB1*09:01:02 | DRB1*15:01:01 |
| Negative | 6.6 | DRB1 | DRB1*08:02:01 | DRB1*15:01:01 |
| Negative | 6.5 | DRB1 | DRB1*10:01:01 | DRB1*14:216 |
| Negative | 5.7 | DRB1 | DRB1*07:01:01 | DRB1*11:04:01 |
| Negative | 5.7 | DRB1 | DRB1*15:02:01 | DRB1*15:02:01 |
| Negative | 5.6 | DRB1 | DRB1*03:01:01 | DRB1*16:01:01 |
| Negative | 5.5 | DRB1 | DRB1*03:01:01 | DRB1*04:05:01 |
| Negative | 4.7 | DRB1 | DRB1*07:01:01 | DRB1*15:03:01 |
| Negative | 4.3 | DRB1 | DRB1*07:01:01 | DRB1*08:01:01 |
| Negative | 3.6 | DRB1 | DRB1*03:01:01 | DRB1*13:01:01 |
